# Delayed Negative Symptom Response To Antipsychotics: An Individual Participant Data Analysis

**DOI:** 10.1101/2025.06.09.25329268

**Authors:** Stefan Leucht, Mathias Harrer, Susan Illing, Stephen Z. Levine

## Abstract

This protocol outlines an individual participant data (IPD) analysis investigating the temporal dynamics of schizophrenia symptoms during antipsychotic treatment. Using harmonized data from six antipsychotic drug trials, we aim to examine weekly trajectories of Positive and Negative Syndrome Scale (PANSS) sub-scores over a 26-week period. Our aim is to determine whether reductions in positive symptoms precede and predict improvements in negative symptoms. To this end, will conduct (i) descriptive analyses, showing the trajectories of five PANSS sub-scores over time; (ii) test potential a “delay” until response in negative symptoms (versus positive symptoms) is reached; and (iii) examine, in exploratory analyses, whether week-to-week improvements in one symptom domain predict subsequent changes in the others, in particular whether improvements in positive symptoms precede changes in negative and cognitive symptoms.

## Background & Aims

The aim of this individual participant data (IPD) analysis is to examine the trajectory of schizophrenia symptoms during antipsychotic treatment. Antipsychotic drugs are believed to only improve positive (delusions, hallucinations), but not the negative symptoms of schizophrenia. However, in randomised, double-blind trials comparing antipsychotic drugs with placebo (Leucht, Arbter et al., 2009) and comparing antipsychotic drugs head-to-head (Leucht, Corves, et al., 2009; Huhn et al., 2019) both positive and negative symptoms improve, even with high-potency, firstgeneration *D*_2_ blockers such as haloperidol. One hypothesis is that the improvement of negative symptoms comes after the improvement of positive symptoms in the following way: patients suffering from hallucinations and delusions are so concerned by their symptoms that they do not interact with other people. When positive symptoms are improved by antipsychotic drugs, these negative symptoms will improve as well. We will harness IPD from six large-scale clinical trials in patients with acute schizophrenia. This study aims to assess whether reductions in positive symptoms precede and are associated with subsequent improvements in negative symptom burden over time.

### Eligibility criteria

Individual participant data (IPD) for this study are drawn from six large-scale randomized trials of antipsychotic treatment (Breier et al., 2005; Colonna et al., 2000; Kahn et al., 2008; Lieberman et al., 2003; Sechter et al., 2002; Tran et al., 1997). These IPD have previously been used in analyses by our group (Leucht et al., 2007; Beitinger et al., 2008; Levine & Leucht, 2012; Levine & Leucht, 2010). Participants are included if they meet the following criteria: (i) a diagnosis of acute schizophrenia (paranoid, disorganized, or undifferentiated type), schizophreniform disorder, or schizoaffective disorder according to DSM-III-R through DSM-5; (ii) received therapeutically active doses of second-generation antipsychotics, as defined by Gardner et al. and McAdams et al.; (iii) were treated with second-generation antipsychotics only. In trials where multiple second-generation antipsychotics were studied, we will select the agent associated with the lowest risk of extrapyramidal side effects, based on findings by Huhn et al. (2019). This is because extrapyramidal symptoms such as parkinsonism and akinesia can mimic negative symptoms, thereby confounding the analysis. For the same reason, first-generation dopamine *D*_2_ receptor antagonists, such as haloperidol, are excluded. Additionally, we will exclude all participants who received (i) placebo or (ii) “pseudo-placebo” treatments, i.e., clearly sub-therapeutic doses of antipsychotics. To ensure comparability of follow-up assessments, and because no substantial further effects of acute antipsychotic treatment are expected beyond six months, we will restrict all analyses to outcome data from the first 26 weeks (6 months) of treatment in our main analysis.

### Study identification & selection

Studies were identified through a search among the randomised-controlled trials for which our team has individual-patient-data .

### Risk of bias assessment

Risk of bias of the included studies will be evaluated using domains of the Cochrane Risk of Bias (RoB 2) tool (Sterne et al., 2019): randomisation process, deviation from the intervention, missing outcome data, measurement of the outcome, and selection of the reported results.

### Data harmonization

In each study, measures of the Positive and Negative Syndrome Scale (PANSS; Kay et al., 1987) will be transformed into five subscores based on the consensus model proposed by Wallwork et al. (2012). This model classifies 20 original PANSS items into five distinct factors: positive, negative, disorganized/concrete, excited, and depressed. Adopting this structure will allow us to represent the multidimensional structure of schizophrenia while maintaining a high level of comparability within the research domain. One otherwise eligible study (Colonna et al., 2000) provided individual item scores only for PANSS items 1 to 14. To approximate the five-factor scores for this study, missing items will be supplemented with corresponding items from the Brief Psychiatric Rating Scale (BPRS; Overall & Gorham, 1962). As a result, two factors will be fully reconstructed, and three remain incomplete, with one missing item each.

### Missing data handling

Primary analyses will be conducted in the observed data. As a sensitivity analysis, missing data will also be addressed using multiple imputation by fully conditional specification (MICE algorithm; Buuren & Groothuis-Oudshoorn, 2011), under the missing-at-random (MAR) assumption. To assess the plausibility of the MAR assumption, exploratory analyses will be conducted, including visualizations of imputed values and examination of missingness patterns. Multilevel imputation models with homoscedastic errors will be used to account for the nesting of measurements within patients and trials. Highly collinear variables and variables with systematically missing data (“structural zeros”) will be excluded from the imputation model. Predictive mean matching will be used as the default imputation method. A total of *m = 25* imputed datasets will be generated. Analysis models will be fitted in each dataset, and parameter estimates will be combined using Rubin’s rules.

### Statistical analyses

To characterize the dataset, we will analyze weekly scheduled visits across the five PANSS subscales using descriptive statistics. For each study and for the pooled data, we will calculate and visualize (i) the weekly mean subscale score and its standard deviation, (ii) the mean absolute change per subscale, (iii) the weekly mean change relative to the baseline values (expressed as a percentage), and (iv) the mean cumulative change from baseline over six months as a percentage of the baseline value.

To examine the possibility of a delayed response in negative symptoms following antipsychotic treatment, we will define a 50% reduction on each Wallwork subscale as a treatment response. For each participant, we will determine the week in which this response criterion is first met, yielding time-to-event data (which may be right-censored). Based on these data, we will calculate the median time-to-response in weeks for each subscale, and test differential response times using multi-level Cox regression models with patients nested in trials, adjusted for baseline symptom severity, age and sex. In addition to within-trial clustering, these models account for the within-patient dependence arising from the five domain-specific response times contributed by each patient, and are weighted using inverse-probability-of-censoring weights (IPCW) to address potentially informative censoring. The 50% threshold is applied to each five-factor Wallwork subscore and computed from raw PANSS scores after subtracting the theoretical minimum score for each sub-scale; note that this cut-off is established primarily for PANSS total score change and is here applied separately to each domain.

To investigate whether improvements in positive symptoms may be statistically associated with improvements in other symptom domains, we will employ path analyses based on structured equation modeling (SEM). Specifically, we will estimate direct, indirect, and total path effects over six months. Additionally, we will conduct path analyses with negative symptoms as the outcome. These path analyses are exploratory, and their estimates are interpreted as statistical associations rather than causal or mediation effects. Following recommendations during peer review, the principal path model was set to a pooled cross-lagged panel model estimated within an SEM framework in long format, including autoregressive and cross-lagged paths and using all weekly time points, with cluster-robust standard errors at the patient level; this model addresses which domains predict week-to-week change in the others. The two-time-point model, using baseline and change to endpoint (at 26 and 6 weeks), is retained as a simpler complementary analysis. Following a reviewer request, we will additionally conduct path analyses with cognitive/disorganization symptoms as the outcome, alongside negative and positive symptoms.

The primary analysis is the multilevel Cox model based on the 50% response threshold in the observed data, comparing the time-to-response of positive versus negative symptoms. All other domain comparisons, alternative sample specifications, the descriptive trajectories, and the path analyses are sensitivity or exploratory analyses; accordingly, no adjustment for multiplicity is made.

### Sensitivity analyses

We will conduct several sensitivity analyses to assess the robustness of our findings. First, where applicable, we will repeat all primary analyses using multiply imputed data (see “missing data handling” section). Second, we will repeat all analyses when only haloperidol arms are excluded due to its pronounced extrapyramidal symptoms risk. Third, we will rerun all analyses using only the subset of trials with outcome data available up to one year (52 weeks). Fourth, we will rerun time-to-response analyses using a 25% symptom reduction as the response criterion for each Wall-work subscale. Fifth, we will also investigate a shorter period of six weeks in the path analyses. Sixth, we will apply cluster-robust variance estimation to the between-domain differences in time-to-response. Seventh, following a reviewer request we will repeat the time-to-response analysis using the original 7-item PANSS positive and negative subscores, in addition to the five-factor scores. Eighth, we will repeat all analyses excluding the one study for which not all PANSS items were available (Colonna et al., 2000).

### Current status

This protocol and statistical analysis plan (SAP) was registered after identifying and harmonizing eligible data, but prior to conducting any statistical analyses. The table below provides an overview of the antipsychotic trials in the current IPD, including study characteristics and eligible sample sizes for the present analysis.

| Study | Medication (Dose) | N (Included) | Inclusion Criteria & Exclusion Criteria | Country | Assessments (Weeks) |
| --- | --- | --- | --- | --- | --- |
| Breier et al., 2005 | Olanzapine 10-20mg | 277 | *Inclusion:* Diagnosis of schizophrenia (DSM-IV); in- and outpatients; BPRS baseline score $\geq 42$, PANSS score $\geq 4$ on at least one positive symptom item and a CGI severity of illness score $\geq 4$.<br>*Exclusion:* Participation in clinical trial of another drug within 1m before study entry; treatment with injectable depot antipsychotic within one treatment cycle before study entry; treatment with clozapine within 7d before enrollment; treatment with olanzapine or ziprasidone within 6m of study start and treatment withdrawal because of clinically important and/or intolerable adverse events or lack of treatment response; congenital long QT syndrome; QTc interval longer than 500 msec, or any ECG abnormality that would confound measurement of the QT interval; substance dependence (DSM-IV) within past month; serious/unstable illness. | Argentina, Austria, Brazil, Germany, Great Britain, Ireland, Mexico, Peru, Puerto Rico, Spain, USA, Venezuela | 0, 1, 2, 3, 4, 5, 6, 8, 10, 13, 16, 20, 24 |
| Colonna et al., 2000 | Amisulpride 200-800mg | 368 | *Inclusion:* Diagnosis of subchronic or chronic schizophrenia with acute exacerbation (DSM-III-R) of the paranoid, disorganised or undifferentiated type; BPRS score $\geq 4$ on at least two positive items.<br>*Exclusion:* Schizophreniform or schizoaffective disorder, mania or excitative depressive states, and severe cardiovascular disease, Parkinson's disease or phaeochromocytoma. | Belgium, France, Great Britain, Hungary, Italy, Latvia, Morocco, Slovenia, Spain | 0, 1, 2, 4, 13, 26 |
| Kahn et al., 2008 | Amisulpride 200–800mg, Olanzapine 5–20mg | 105 | *Inclusion:* Diagnosis of schizophrenia, schizophreniform disorder, or schizoaffective disorder (DSM-IV) confirmed by the MINI plus<br>*Exclusion:* More than 2y since the onset of positive symptoms; use of any antipsychotic drug for more than 2w in previous year, or for 6w at any time; known intolerance to one of the study drugs; contraindications for any of the study drugs. | Austria, Belgium, Bulgaria, Czech Republic, France, Germany, Israel, Italy, Netherlands, Poland, Romania, Spain, Sweden, Switzerland | 0, 2, 4, 6, 8, 12, 26 |
| Lieberman et al., 2003 | Olanzapine 5-10mg | 131 | *Inclusion:* Diagnosis of schizophrenia, schizophreniform disorder, or schizoaffective disorder (DSM-IV); onset of psychotic symptoms before age 35 years; psychotic symptoms (delusions, hallucinations, thought disorder, and grossly bizarre behavior) for at least 1m but not more than 60m, PANSS score $\geq 4$ on at least two psychosis items or score $\geq 5$ on one psychosis item; CGI severity score $\geq 4$; required treatment with antipsychotic drugs on a clinical basis<br>*Exclusion:* Previously antipsychotic drug treatment for more than 16 cumulative weeks; treatment with clozapine at anytime in lifetime, or treatment with an injectable depot neuroleptic within less than three dosing intervals before study entry; pregnancy or lactation, or no adequate use of contraception; serious, unstable medical illnesses or findings from a medical examination that suggested a contraindication to antipsychotic drug treatment; history of allergic or severe adverse reactions to study medications; diagnosis of substance dependence (DSM-IV) (except caffeine and nicotine) within 1m before first visit; serious suicidal risk; required treatment with anticonvulsants, benzodiazepines (except as allowed for agitation and control of extrapyramidal signs), antidepressants, psychostimulants, or other antipsychotic drugs concurrently with study medications beyond those permitted as concomitant treatments; contraindications for neuroimaging per current regulations of the local regulatory agency; history of any DSM-IV psychotic disorder with recovery; premorbid IQ of $\leq 70$; ECT within 1m before study entry | Canada, Great Britain, Netherlands, USA | 0, 1, 2, 3, 4, 5, 6, 8, 10, 12, 16, 20, 24 |
| Sechter et al., 2002 | Amisulpride 400-1000mg | 152 | *Inclusion:* Diagnosis of chronic schizophrenia (disease duration of at least 2y) of the paranoid, disorganized, undifferentiated, or residual type (DSM-IV); PANSS total score of 60-120 and recent deterioration of clinical state needing a change in treatment; in- or outpatients<br>*Exclusion:* Predominant negative symptoms (3 or more items $\geq 4$ on the negative PANSS subscale and less than 3 items $\geq 4$ on the positive PANSS); symptoms of substance-related withdrawal; cardiovascular disease, severe respiratory, hematologic or endocrinologic diseases, Parkinson's disease, pheochromocytoma; significant abnormal laboratory tests, or a history of allergy or hypersensitivity to risperidone or benzamides; current treatment with amisulpride or risperidone; no response to at least two well-validated antipsychotics; received lithium-based therapy in previous 10d; received long-acting antipsychotics (if the usual interval between two injections had not elapsed before inclusion); pregnancy or lactation, and women of childbearing age not using adequate contraception | Austria, Belgium, Estonia, France, Germany, Hungary, Latvia, Netherlands, Slovenia | 0, 1, 2, 3, 4, 6, 8, 13, 17, 21, 26 |
| Tran et al., 1997 | Olanzapine<br>10-20mg | 172 | *Inclusion:* Diagnosis of schizophrenia, schizophreniform disorder, or schizoaffective disorder (DSM-IV); BPRS score $\geq 42$; in- or outpatients<br>*Exclusion:* Comorbid or other recent major axis I disorders; pregnancy or lactation; serious medical illnesses in which pharmacotherapy posed a substantial risk or confounded diagnosis; failed to show at least minimal clinical response with at least three antipsychotics for at least 6w. | Belgium, France,<br>Germany, Great Britain,<br>Netherlands,<br>South Africa, Spain,<br>Switzerland | 0, 1, 2, 3, 4,<br>5, 6, 7, 8,<br>12, 16, 20,<br>24 |

The following decisions were made for drug choice:

- In Breier et al. (2005), Kahn et al. (2008) and Tran et al. (1997), we chose olanzapine and amisulpride rather than ziprasidone or risperidone because of their lower EPS risk according to Huhn et al. (2019).
- For the same reason, amisulpride was preferred to risperidone in Sechter et al. (2002).
- Olanzapine was preferred to quetiapine in Kahn et al. (2008), because in Huhn et al. (2019) the EPS risk is the same, but olanzapine has consistently been shown to be a more efficacious drug (Huhn et al. 2019, Leucht et al., 2013; Taipale et al., 2018).
- Haloperidol arms were generally excluded (see “Eligibility criteria”).

### Explanation of protocol changes

Mainly due to reviewer suggestions there were some changes of the original protocol which we present here

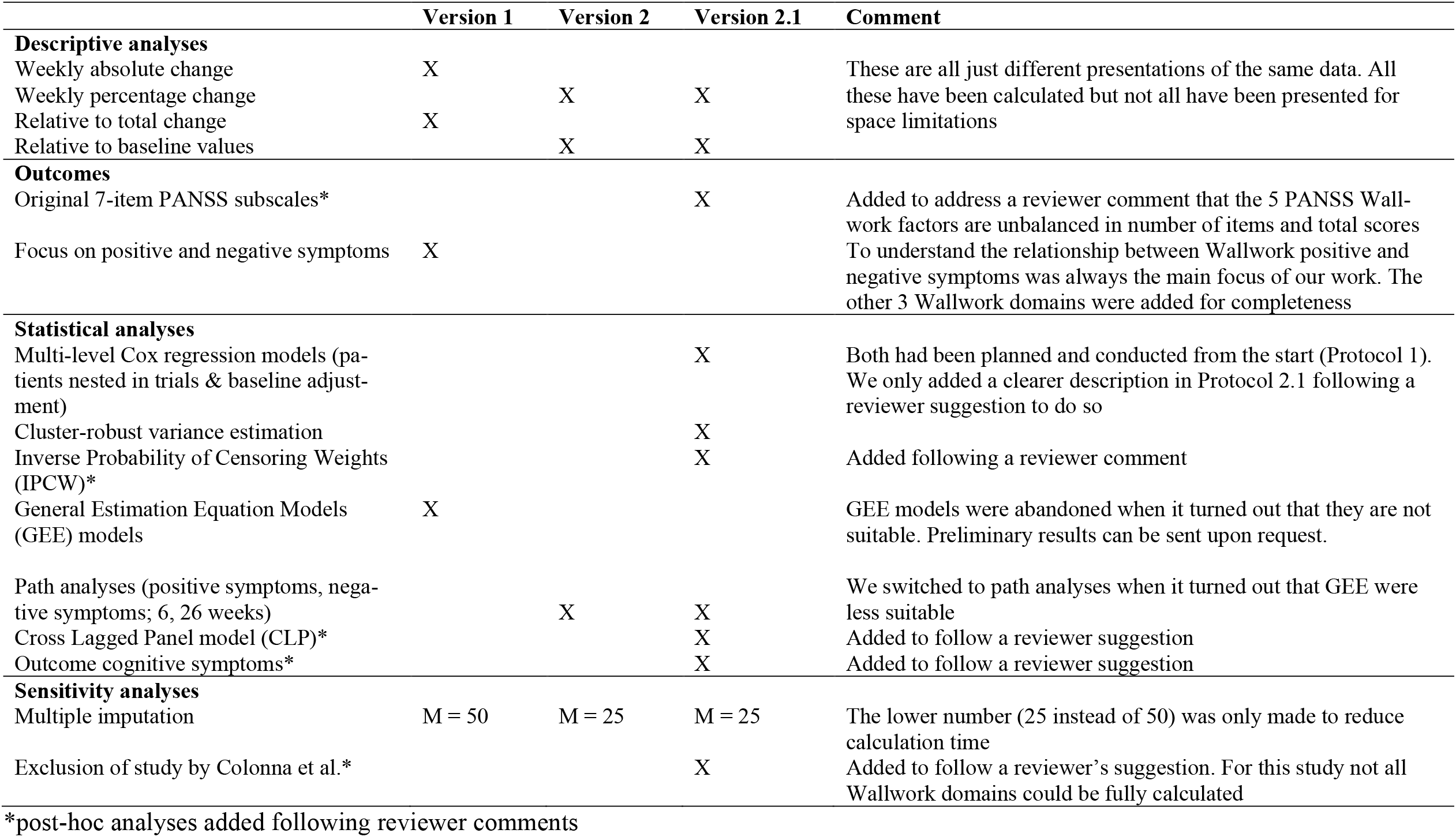

*post-hoc analyses added following reviewer comments

### Amendments in version 2.2

In Version 2.2 an overview and explanation of protocol changes was added. The table compares the different protocol versions and highlights post-hoc analyses following reviewer comments.

## Data Availability

Aggregate-level data of the present study are available upon reasonable request to the authors. Patient-level individual participant data (IPD) was analyzed locally using the EBMPP Trial Data Warehouse hosted at TU Munich but cannot be shared with third parties due to data confidentiality.

